# Regional differences in time off work after injury: a comparison of Australian states and territories within a single workers’ compensation system

**DOI:** 10.1101/2020.07.23.20160416

**Authors:** Tyler J Lane, Luke Sheehan, Shannon Gray, Alex Collie

## Abstract

**Background:** Time off work after workplace injury varies by compensation system. While often attributed to features of the compensation system, unaccounted regional factors may drive much of the effect. In this study, we compare disability durations by state and territory of residence within a single national workers’ compensation system. Large differences would indicate that factors other than compensation system settings are responsible for system effects observed in previous studies.

**Methods:** We applied crude and adjusted Cox proportional hazards models to compare disability durations by state and territory of residence. Confounders included factors known to influence disability duration. Durations were left-censored at two weeks and right-censored at 104 weeks.

**Results:** We analysed *N* = 38,686 claims. In both crude and adjusted models, three of the seven states and territories significantly differed from the reference group, New South Wales. However, two of the three were different between crude and adjusted models. Regional effects were relatively small compared to other factors including insurer type, age, and type of injury.

**Conclusions:** Regional factors influence disability duration, which persist with adjustment for demographic, work, insurer type, and injury confounders. However, the effects are inconsistently significant and fairly small, especially when compared to the effect of confounders and system effects found in previous studies. Regional factors likely only account for a small share of the difference in disability duration between compensation systems.

## Introduction

Workers’ compensation insurance covers part or all of the wages of people who are off work due to occupational injury or illness. The amount of compensated time off work, referred to here as disability duration, differs between compensation systems, and persists with adjustment for known confounders.^1–3^ We previously suggested that differences between Australia’s state, territory, and Commonwealth systems are attributable to compensation system factors such as policy, practice, and design.^2^ However, attributing residual differences between groups to a specific yet unmeasured factor risks the residual fallacy.^4^ Any number of unmeasured or imperfectly measured confounders or other factors may account for the difference, and in practice, it is impossible to account for all sources of confounding.

The residual fallacy problem is amplified within compensation systems, which are often regionally-bound to national and subnational political entities. In Australia, structural and policy differences that are independent of workers’ compensation yet vary between states and territories may influence disability duration. For instance, the healthcare system is recognised as one of the four main domains of influence over work disability and return to work.^5^ While Australia has a universal health insurance system, states and territories are responsible for running public hospitals and regulating private hospitals.^6,7^ While all work health and safety regulators (excluding Victoria and Western Australia) have adopted the Commonwealth’s model Work Health and Safety Laws,^8^ annual work injury rates vary considerably, from 36/1000 workers in South Australia to 57/1000 in Australian Capital Territory.^9^ There are considerable socio-economic differences as well, which are predictive of time off work.^10,11^ In the Australian Capital Territory, 55% of residents live within the most advantaged quintile of postcodes, compared to 4.6% of Tasmanian residents.^12^ Residents of rural and remote areas have substantially longer disability durations;^13^ some Australian states have less than 1% of their population in remote areas (New South Wales, Victoria, Queensland, Australian Capital Territory), compared to 6% in Western Australia and 40% in the Northern Territory.^14^

Many of these can be statistical adjusted for, but often only imprecisely. Other, more abstract factors like cultural differences are also predictive of disability duration and vary regionally.^11^

Differentiating regional and system effects is essential for efforts to reduce disability durations. System effects due to policy, practice, and design features are amenable to modification, whereas regional factors like culture and economics may be less modifiable.

In this study, we test for regional effects on disability duration in order to improve understanding of system effects. Should regional effects be comparable to the system effects we previously observed,^2^ it would suggest that little of the difference between compensation systems is due to system factors. Conversely, if regional effects are substantially smaller than system effects, it would suggest differences are mostly due to system factors. We take advantage of the unique arrangement of workers’ compensation in Australia, where there is a national scheme for the Commonwealth government and multi-jurisdictional employers. This allows us to test for regional effects (i.e., state and territory of residence) while removing differences attributable to system factors, and to compare them to previously observed system effects.

## Methods

### Setting

Comcare is the Australian Commonwealth government’s workers’ compensation regulator and insurer. There is only a limited range of employers within the scheme: the Commonwealth government, whose workforce is insured by the scheme, and multi-jurisdictional employers, whom Comcare issues self-insurance licenses to. As of 30 September 2018, Comcare insured 395,000 workers and regulated coverage of 188,000 workers from self-insured organisations.^15^ Both scheme and self-insured employers must adhere to the same workers’ compensation legislation. Comcare also regulates work health and safety under the Work Health and Safety Act 2011 with the exception of self-insured employers licensed after 2011, who are subject to state and territory regulators.^16^

### Data

Claims data are taken from *National Dataset for Compensation-based Statistics*, an amalgamation of administrative workers’ compensation claims data from each Australian workers’ compensation system that was designed for their integration and comparison.^17^ These are supplemented with the Index of Relative Socio-economic Advantage and Disadvantage (IRSAD)^18^ and the Australian Statistical Geography Standard (ASGS)^19^ which rank postcodes on socio-economic factors and remoteness.

### Inclusion and exclusion criteria

Inclusion criteria were accepted claims lodged in the Comcare system between July 2003 and June 2015, having been compensated for time off work, and being aged 15-80 years at the time of injury. Only claims with at least two weeks of compensated time loss were included, which makes the findings comparable to our previous system effects study.^2^

### Exposures, outcomes, and confounders

Main exposures were state or territory of residence. The outcome was cumulative compensated time off work, matching the operationalisation applied in our previous study of system differences.^2^ Analyses were adjusted for confounders, defined as factors known or hypothesised to affect disability duration and to vary across regions. These included age group (15-24, 35-44, 45-54, 55+ years), sex, socioeconomic advantage and disadvantage of residential postcode (most advantaged quintile, most disadvantaged quintile, and middle three quintiles based on the IRSAD^18^), urban location (major city, other, based on ASGS^19^), full/part-time hours (dichotomised at 35 hours), industry of employer (Australia New Zealand Standard Industrial Classification^20^), occupation (Australia and New Zealand Standard Classification of Occupations^21^), and injury type (Type of Occurrence Classification 3.1,^22^ with adapted groupings of fracture, musculoskeletal, neurological, mental health, other trauma, and disease^23^). Given some small values within some industry groups, we combined those with under 200 cases into an “other” category.

### Analysis

We conducted Cox proportional hazards survival analyses to compare disability durations by state and territory of residence. Hazard ratios above one indicated shorter disability durations. Disability durations were right-censored at 104 weeks. New South Wales served as the reference as it was the largest state/territory of residence. The effect of state and territory of residence was evaluated in both crude and adjusted models. Adjusted survival curves were plotted to illustrate differences across states, which were log-transformed to account for the logarithmic decay pattern of disability durations (illustrated in Figure 1) and to increase the visibility of differences later in the process.

**Figure 1.**
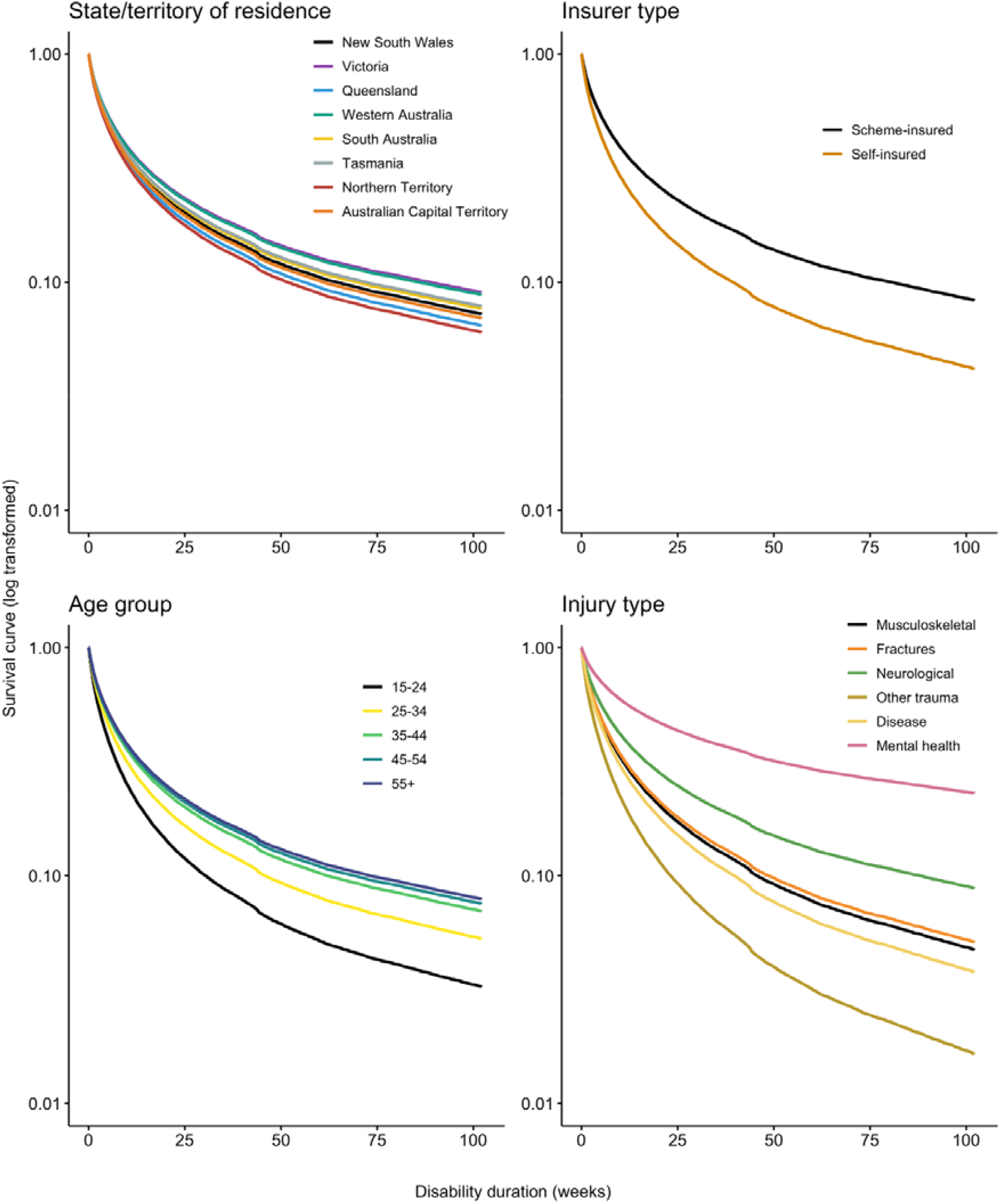
Adjusted Cox survival curves for disability duration by state/territory of residence (regional), insurer type, age group, and injury type; reference groups are in black

We applied multiple imputation to account for missing data. The number of imputations was proportional to the number of cases with missing data (3.6% cases with missing data, therefore 4 imputations). Claims without postcode data or that could not be matched to a state or territory of residence data (*N* = 49, 0.1%) were excluded rather than imputed.

An early version of the analysis plan was hosted on the Open Science Framework.^24^ However, the final design varies so much from the original that we do not consider it fair to consider the study pre-registered. We highlight this for transparency. Analytical code are archived on a Bridges repository.^25^ We are unable to share the data given their sensitivity as case-level claims data.

### Statistical software

Analyses are conducted in R^26^ using RStudio^27^ with the following packages: *broom*,^28^ *cowplot*,^29^ *gridExtra*,^30^ *Epi*,^31^ *janitor*,^32^ *lubridate*,^33^ *mice*,^34^ *naniar*,^35^ *see*,^36^ *survival*,^37^ *survminer*,^38^ *summarytools*,^39^ *tidyverse*,^40^ and *zoo*.^41^

## Results

After applying inclusion/exclusion criteria, we retained *N* = 38,686 claim records for analysis. Population descriptives are presented along with model outputs in Table 1. In the crude model, three of the seven states and territories had hazard ratios that were significantly smaller than the reference group (New South Wales), denoting longer disability durations: Victoria (0.94; 95% confidence interval: 0.91-0.97), Tasmania (0.83; 0.77-0.89), and the Australian Capital Territory (0.88; 0.85-0.91). However, with the inclusion of confounders in adjusted analysis, Tasmania and Australian Capital Territory attenuated to non-significance, while Western Australia’s hazard ratio became significantly smaller (0.91; 0.87-0.95), denoting longer disability durations, and Queensland’s hazard ratio became significantly bigger (1.06; 1.02-1.09), denoting shorter disability durations. Victoria’s hazard ratio remained significantly smaller in the adjusted model (0.90; 0.87-0.93). Adjusted survival curves by state and territory of residence are presented in Figure 1.

**Table 1.** Population descriptive statistics and Cox proportional hazard model results, with significant effects at p ≤ .05 in bold

| Variable | Descriptives - N (%) | Model outputs - Hazard Ratio (95% CI) |  |
| --- | --- | --- | --- |
|  |  | Crude | Adjusted |
| Region – state or territory of residence |  |  |  |
| New South Wales | 11,617 (30.1%) | Ref | Ref |
| Victoria | 7,595 (19.7%) | **0.94 (0.91-0.97)** | **0.90 (0.87-0.93)** |
| Queensland | 5,414 (14.0%) | 1.01 (0.98-1.04) | **1.06 (1.02-1.09)** |
| Western Australia | 2,781 (7.2%) | 0.98 (0.94-1.02) | **0.91 (0.87-0.95)** |
| South Australia | 1,977 (5.1%) | 0.96 (0.91-1.01) | 0.97 (0.93-1.02) |
| Tasmania | 764 (2.0%) | **0.83 (0.77-0.89)** | 0.96 (0.89-1.04) |
| Northern Territory | 398 (1.0%) | 0.96 (0.87-1.07) | 1.09 (0.98-1.21) |
| Australian Capital Territory | 8,091 (20.9%) | **0.88 (0.85-0.91)** | 1.02 (0.98-1.06) |
| Missing | 49 (0.1%) |  |  |
| Self-insured employer | 15,722 (40.6%) |  | **1.34 (1.28-1.41)** |
| Month of claim lodgement (continuous variable) | - |  | **0.98 (0.97-0.98)** |
| Female | 18,026 (46.6%) |  | **0.93 (0.90-0.95)** |
| Age at time of accident (years) |  |  |  |
| 15-24 | 1,086 (2.8%) |  | Ref |
| 25-34 | 6,211 (16.1%) |  | **0.82 (0.77-0.88)** |
| 35-44 | 1,063 (28.6%) |  | **0.73 (0.69-0.78)** |
| 45-54 | 13,582 (35.1%) |  | **0.71 (0.66-0.75)** |
| 55+ | 6,744 (17.4%) |  | **0.69 (0.65-0.74)** |
| Part time employment (<35 hours per week) | 6,089 (15.7%) |  |  |
| Industry |  |  |  |
| Public administration & safety | 16005 (42.7%) |  | Ref |
| Construction | 623 (1.7%) |  | **0.79 (0.71-0.87)** |
| Education & training | 1410 (3.8%) |  | 1.00 (0.94-1.06) |
| Electricity, gas, water, & waste services | 789 (2.1%) |  | 1.01 (0.93-1.10) |
| Financial & insurance services | 1355 (3.6%) |  | 1.05 (0.99-1.13) |
| Health care & social services | 1729 (4.6%) |  | **0.76 (0.72-0.81)** |
| Information media & telecommunications | 2373 (6.3%) |  | 0.96 (0.90-1.02) |
| Manufacturing | 301 (0.8%) |  | **1.24 (1.10-1.41)** |
| Professional, scientific, & technical services | 874 (2.3%) |  | 1.02 (0.94-1.10) |
| Transport, postal, & warehousing | 11658 (31.1%) |  | 1.00 (0.95-1.05) |
| Other | 342 (0.9%) |  | 0.92 (0.83-1.03) |
| Missing | 1,227 (3.2%) |  |  |
| Occupation |  |  |  |
| Clerical & administrative workers | 20,361 (52.6%) |  | Ref |
| Managers | 2,115 (5.5%) |  | 1.03 (0.98-1.08) |
| Professionals | 3,326 (8.6%) |  | **1.05 (1.01-1.10)** |
| Technicians & trades workers | 3,318 (8.6%) |  | **1.05 (1.00-1.10)** |
| Community & personal service workers | 3,042 (7.9%) |  | 0.97 (0.93-1.01) |
| Sales workers | 674 (1.7%) |  | **1.10 (1.02-1.19)** |
| Machinery operators & drivers | 4,419 (11.4%) |  | **0.94 (0.90-0.97)** |
| Labourers | 1,431 (3.7%) |  | **0.92 (0.87-0.98)** |
| Urban |  |  |  |
| Major city | 31,382 (81.5%) |  | Ref |
| Other | 7,129 (18.5%) |  | **0.94 (0.91-0.97)** |
| Missing | 175 (0.5%) |  |  |
| Postcode socio-economic status |  |  |  |
| Bottom quintile | 4,098 (10.6%) |  | 0.97 (0.93-1.00) |
| Middle quintile | 19,553 (50.8%) |  | Ref |
| Top quintile | 14,865 (38.6%) |  | **1.05 (1.02-1.08)** |
| Missing | 170 (0.4%) |  |  |
| Injury type |  |  |  |
| Musculoskeletal condition | 24,992 (64.6%) |  | Ref |
| Fractures | 3,078 (8.0%) |  | 0.97 (0.94-1.01) |
| Neurological | 958 (2.5%) |  | **0.79 (0.74-0.84)** |
| Mental health | 3,008 (7.8%) |  | **0.47 (0.45-0.48)** |
| Other trauma | 1,523 (3.9%) |  | **1.37 (1.32-1.43)** |
| Disease | 5,127 (13.2%) |  | **1.08 (1.02-1.14)** |
| Total | 38,686 |  |  |

While not main exposures, a number of confounders had consistently larger effects than region and are worth highlighting. We have plotted these along with regional effects in Figure 1 to illustrate relative magnitudes. Claims from self-insured employers had larger hazard ratios (1.34; 1.28-1.41). Age followed a stepped effect, with hazard ratios decreasing and plateauing with age. Relative to musculoskeletal conditions, disability durations across all injury types were significantly different except for fractures. Mental health conditions had the smallest hazard ratios (0.47; 0.45-0.48) while other traumas were the biggest (1.37; 1.32-1.43).

Notably, survival curves in Figure 1 exhibit a slight step shift just before the 50 week mark. This corresponds with a step-down in wage replacement from 100% to 75% of pre-injury earnings occurring at 45 weeks in Comcare, which we previously found had a small but significant effect on disability duration.^42^

## Discussion

We previously found substantial differences in disability durations between Australian workers’ compensation systems.^2^ However, these systems correspond to states and territory geographic boundaries, potentially confounding analyses with regional differences. The aim of this study was to test for regional effects on disability duration within a single workers’ compensation system, thereby keeping compensation system settings constant.

We found some evidence of regional effects on disability duration. Of the seven states and territories compared to New South Wales (the reference region), three differed significantly. However, two of the three changed between crude and adjusted models. Victoria had significantly longer durations in both, though Tasmania and Australian Capital Territory were only significantly different in the crude model while Queensland and Western Australia were only significantly different after adjustment for confounders. While our aim was not to identify the regions that performed better or worse, differences between crude and adjusted models are a useful consistency check. This may reflect a problem of big datasets like the one used in this study, where inconsequential effects emerge as statistically significant, giving them artificial importance.^43^ In this case, slight changes in small effects between crude and adjusted models could lead individual coefficients to slip in and out of significance without a meaningful change in effect.

The magnitude of regional effects may offer better insight as to their importance. In Figure 2, we present adjusted effect estimates with 95% confidence intervals from the current study, along with system effects from our previous study, which compared disability durations across state and territory compensation systems.^2^ With the exception of Comcare, which is included as a separate compensation system, these correspond to the states and territories of residence in the current study. We replicated the previous study’s analysis, updating the confounder approach to match that in the current study, though retain the original restriciton of claims to those lodged in 2010.

**Figure 2.**
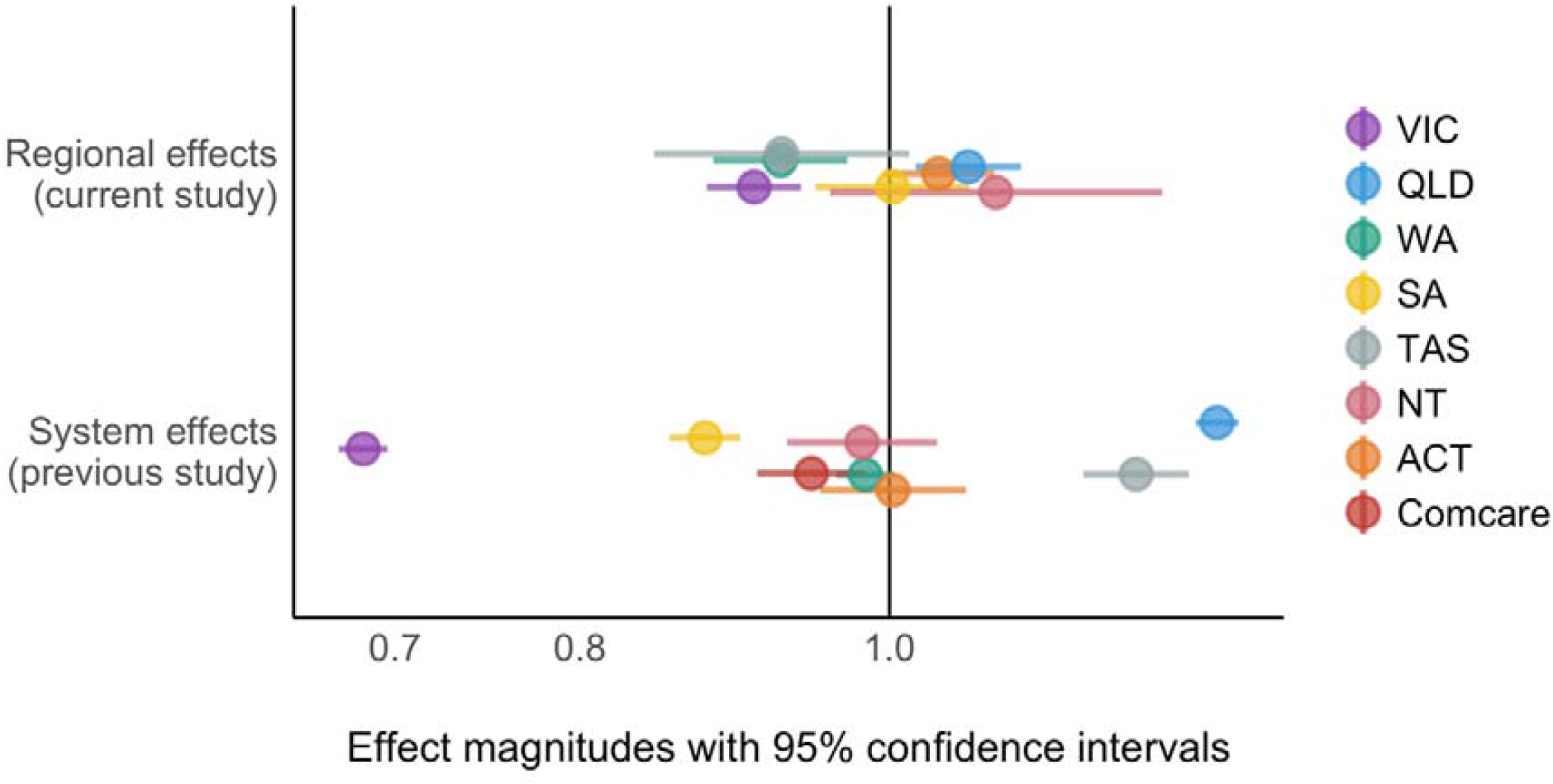
Comparison of effect magnitudes between current study and previous system comparison study^2^

As illustrated in Figure 2, effect variance is greater between systems than regions. This suggests that a substantial proportion of the variance between compensation systems is attributable to the systems themselves. In other words, system features such as policy, practice, and design have a greater impact on disability duration than regional factors. This is encouraging because many system features can be modified to reduce disability durations, whether through legislation or a change in practice. However, this leaves unanswered the question of which system features actually influence disability duration, and which of those are most important. There is a body of research linking a number of system features to disability duration including experience of the claims process and delays in claim processing,^44,45^ fee schedules, healthcare provider choice, retroactive periods,^46,47^ and rate of compensation.^42,48,49^ However, modifications such as legislative change must be carefully considered as they may not work as intended, and in some cases may even have unintended negative consequences.^50^

There is one important caveat to the comparison of system and regional effect magnitudes. Comcare claims come from only two types of employers: scheme-insured Commonwealth and self-insured multijurisdictional companies. This is quite different from other compensation systems, and likely results in greater homogeneity and potentially less variability in disability duration.

Several confounders had a greater effect on disability duration than region. Notably, self-insured claims had much shorter disability durations than scheme-insured claims, which aligns with previous findings.^51,52^ Self-insurers have been found to process claim applications faster,^51^ which, as noted above, is predictive of shorter disability durations.^44,45^ As both scheme and self-insured employers are subject to the same workers’ compensation regulations, this shows the importance of non-policy factors like employer practices and highlights the variability within compensation systems. However, as noted elsewhere, scheme and self-insured employers are very different from each other in Comcare; the former is the Commonwealth government, while the latter are multi-jurisdictional companies. We previously found that claims from government employers are similar to other industries, if not shorter,^53^ which means the difference between scheme and self-insurers could be even larger. Future compensation systems research would do well to take scheme and self-insurance arrangements into account. Claimant age and injury both had larger effects on disability duration than region. However, this sheds little light on the effect of region and is well-established in the evidence-base,^2,54^ so we do not expand on this further.

### Strengths and limitations

This study used population-level national claims data from a single workers’ compensation system to isolate regional effects from system effects. Analyses included almost 40,000 claims lodged over a 12-year period and adjusted for a wide range of confounders. However, large datasets can sometimes flag inconsequential effects as significant and exaggerate their importance.

Disability duration, as derived from cumulative compensated time loss, underestimates true time loss.^55^ The end of compensation does not necessarily indicate return to work, as claimants may retire or move to alternative income support systems.^56^ Claims from Comcare differ from other compensation systems by virtue of the types of employers covered, which limits the ability to compare intra-jurisdictional regional effects to inter-jurisdictional system effects.

## Conclusions

After adjustment for confounders, the state and territory of residence significantly influenced disability duration following workplace injury. However, the effect was inconsistently significant and relatively small compared to factors like insurer and injury type and age of the claimant. It was also smaller than the effect of the compenensation system in which the claim was lodged. This suggests that much of the difference between compensation systems is in fact attributable to the system factors such as policy, practice, and design. This is encouraging for researchers and policymakers as system factors can often be modified to improve outcomes.

## Institution and Ethics approval and informed consent

This study received ethics approval from the Monash University Human Research Ethics Committee (CF14/2995 – 2014001663).

## Data Availability

We are unable to share the data given their sensitivity as case-level claims data.

https://doi.org/10.26180/5f1933f8dcb6b

## Data statement

This report uses data supplied by Safe Work Australia and has been compiled in collaboration with state, territory, and Commonweatlh workers’ compensation regulators. The views expressed are the responsibility of the authors and are not necessarily the views of Safe Work Australia or the state, territory, and Commonwealth workers’ compensation regulators.

## References

1. Anema, J. R. et al.. Can cross country differences in return-to-work after chronic occupational back pain be explained? An exploratory analysis on disability policies in a six country cohort study. J. Occup. Rehabil. 19, 419–426 (2009).

2. Collie, A., Lane, T. J., Hassani-Mahmooei, B., Thompson, J. & McLeod, C. B. Does time off work after injury vary by jurisdiction? A comparative study of eight Australian workers’ compensation systems. BMJ Open 6, e010910 (2016).

3. Gray, S. E. & Collie, A. Comparing time off work after work-related mental health conditions across Australian workers’ compensation systems: a retrospective cohort study. Psychiatry, Psychol. Law 8719, (2018).

4. Sowell, T. The Vision of the Anointed: Self-Congratulation as a Basis for Social Policy. (Basic Books, 1995).

5. Loisel, P. et al.. Disability prevention: New paradigm for the management of occupational back pain. Dis. Manag. Heal. Outcomes 9, 351–360 (2001).

6. Krassnitzer, L. The public health sector and Medicare. in Understanding the Australian Health Care System (eds. Willis, E., Reynolds, L. & Trudy Rudge) 18–36 (Elsevier Australia, 2020).

7. Collyer, F., Willis, K. & Keleher, H. The private health sector and private health insurance. in Understanding the Australian Health Care System (eds. Willis, E., Reynolds, L. & Rudge, T.) 37–52 (Elsevier Australia, 2020).

8. Safe Work Australia. Model WHS Laws. https://www.safeworkaustralia.gov.au/law-and-regulation/model-whs-laws.

9. Australian Bureau of Statistics. 6324.0 Work-Related Injuries, Australia, Jul 2017 to Jun 2018. https://www.abs.gov.au/ausstats/abs@.nsf/mf/6324.0 (2018).

10. Shraim, M., Cifuentes, M., Willetts, J. L., Marucci-Wellman, H. R. & Pransky, G. S. Regional socioeconomic disparities in outcomes for workers with low back pain in the United States. Am. J. Ind. Med. 60, 472–483 (2017).

11. Gaines, B., Besen, E. & Pransky, G. S. The influence of geographic variation in socio-cultural factors on length of work disability. Disabil. Health J. 10, 308–319 (2017).

12. Australian Bureau of Statistics. 2071.0 - Census of Population and Housing: Reflecting Australia - Stories from the Census, 2016. https://www.abs.gov.au/ausstats/abs@.nsf/Lookup/bySubject/2071.0~2016~MainFeatures~Socio-EconomicAdvantageandDisadvantage~123 (2018).

13. Macpherson, R. A. et al.. Urban-rural Differences in the Duration of Injury-related Work Disability in Six Canadian Provinces. J. Occup. Environ. Med. 62, e200–e207 (2020).

14. Australian Bureau of Statistics. 3218.0 - Regional Population Growth, Australia, 2018-19. https://www.abs.gov.au/AUSSTATS/abs@.nsf/Lookup/3218.0Main+Features12018-19?OpenDocument (2020).

15. Safe Work Australia. Comparison of Workers’ Compensation Arrangements in Australia and New Zealand. https://www.safeworkaustralia.gov.au/doc/comparison-workers-compensation-arrangements-australia-and-new-zealand-2019 (2020).

16. Commonwealth of Australia. Work Health and Safety Act 2011. (2011).

17. Safe Work Australia. National Data Set for Compensation-based Statistics Third Edition. (2004).

18. Australian Bureau of Statistics. 2033.0.55.001 - Census of Population and Housing: Socio - Economic Indexes for Areas (SEIFA), Australia, 2016. (2018).

19. Australian Bureau of Statistics. Remoteness Structure. (2018).

20. Australian Bureau of Statistics. 1292.0 - Australian and New Zealand Standard Industrial Classification (ANZSIC), 2006 (Revision 2.0). (2013).

21. Australian Bureau of Statistics. ANZSCO: Australian and New Zealand Standard Classification of Occupations. 854 (2006).

22. Australian Safety and Compensation Council. Type of Occurrence Classification System 3rd Edition Revision 1. (2008).

23. Collie, A. & Gray, S. E. ComPARE - Approach to Injury and Condition Coding. (2016).

24. Lane, T. J. Does time off work after injury vary by region? A comparative study of disability durations across Australian states and territories within a single workers’ compensation system. Open Science Framework https://osf.io/2umzb/ (2020) doi:10.17605/OSF.IO/2UMZB.

25. Lane, T. J., Sheehan, L. R., Gray, S. E. & Collie, A. Regional differences in time off work after injury: a comparison of Australian states and territories within a single workers’ compensation system. Bridges https://doi.org/10.26180/5f1933f8dcb6b (2020) doi:10.26180/5f1933f8dcb6b.

26. R Core Team. R: A language and environment for statistical computing. (2020).

27. RStudio Team. RStudio: Integrated Development for R. (2019).

28. Robinson, D. & Hayes, A. broom: Convert Statistical Analysis Objects into Tidy Tibbles. (2020).

29. Wilke, C. O. cowplot: Streamlined Plot Theme and Plot Annotations for ‘ggplot2’. (2019).

30. Auguie, B. gridExtra: Miscellaneous Functions for ‘Grid’ Graphics. (2017).

31. Carstensen, B., Plummer, M., Laara, E. & Hills, M. Epi: A Package for Statistical Analysis in Epidemiology. (2019).

32. Firke, S. janitor: Simple Tools for Examining and Cleaning Dirty Data. (2020).

33. Grolemund, G. & Wickham, H. Dates and Times Made Easy with lubridate. J. Stat. Softw. 40, 1–25 (2011).

34. van Buuren, S. & Groothuis-Oudshoorn, K. mice: Multivariate Imputation by Chained Equations in R. J. Stat. Softw. 45, 1–67 (2011).

35. Tierney, N., Cook, D., McBain, M. & Fay, C. naniar: Data Structures, Summaries, and Visualisations for Missing Data. (2020).

36. Lüdecke, D., Makowski, D., Waggoner, P. & Ben-Shachar, M. S. see: Visualisation Toolbox for ‘easystats’ and Extra Geoms, Themes, and Color Palettes for ‘ggplot2’. (2019).

37. Therneau, T. A Package for Survival Analysis in S. (2015).

38. Kassambara, A. & Kosinski, M. survminer: Drawing Survival Curves using ‘ggplot2’. (2018).

39. Comtois, D. summarytools: Tools to Quickly and Neatly Summarize Data. (2020).

40. Wickham, H. tidyverse: Easily Install and Load the ‘Tidyverse’. (2017).

41. Zeileis, A. & Grothendieck, G. zoo: S3 Infrastructure for Regular and Irregular Time Series. J. Stat. Softw. 14, 1–27 (2005).

42. Lane, T. J., Sheehan, L. R., Gray, S. E., Beck, D. & Collie, A. Step-downs reduce workers compensation payments to encourage return to work. Are they effective? Occup. Environ. Med. (2020) doi:10.1136/oemed-2019-106325.

43. Kaplan, R. M., Chambers, D. A. & Glasgow, R. E. Big data and large sample size: A cautionary note on the potential for bias. Clin. Transl. Sci. 7, 342–346 (2014).

44. Gray, S. E., Lane, T. J., Sheehan, L. R. & Collie, A. Association between workers’ compensation claim processing times and work disability duration: Analysis of population level claims data. Health Policy (New. York). 123, 982–991 (2019).

45. Collie, A., Sheehan, L. R., Lane, T. J., Gray, S. E. & Grant, G. M. Injured worker experiences of insurance claim processes and return to work: a national, cross-sectional study. BMC Public Health 19, 1–12 (2019).

46. Shraim, M., Cifuentes, M., Willetts, J. L., Marucci-Wellman, H. R. & Pransky, G. S. Length of disability and medical costs in low back pain: Do state workers’ compensation policies make a difference? J. Occup. Environ. Med. 57, 1275–83 (2015).

47. Shraim, M., Cifuentes, M., Willetts, J. L., Marucci-Wellman, H. R. & Pransky, G. Why does the adverse effect of inappropriate MRI for LBP vary by geographic location? An exploratory analysis. BMC Musculoskelet. Disord. 20, 1–11 (2019).

48. Hansen, B., Nguyen, T. & Waddell, G. R. Benefit Generosity and Injury Duration: Quasi - Experimental Evidence from Regression Kinks. http://ftp.iza.org/dp10621.pdf (2017).

49. Lane, T. J., Gray, S. E., Sheehan, L. R. & Collie, A. Increased benefit generosity and the impact on workers’ compensation claiming behavior: an interrupted time series study in Victoria, Australia. J. Occup. Environ. Med. 61, e82–e90 (2019).

50. Lane, T. J., Gray, S. E., Hassani-Mahmooei, B. & Collie, A. Effectiveness of employer financial incentives in reducing time to report worker injury: an interrupted time series study of two Australian workers’ compensation jurisdictions. BMC Public Health 18, 1–10 (2018).

51. Sheehan, L. R., Lane, T. J., Gray, S. E., Beck, D. & Collie, A. Comparison of Return to Work Practices and Outcomes in Self-Insured and Scheme-Insured Organisations. https://www.monash.edu/__data/assets/pdf_file/0008/1617281/COMPARE_Self-insurers_with-DOI.pdf (2018) doi:10.26180/5c354a7d235e1.

52. Seabury, S. A., McLaren, C. F., Robert, R., Frank, N. & John, M. Workers’ Compensation Experience Rating and Return to Work. Policy Pract. Heal. Saf. 10, 97–115 (2012).

53. Sheehan, L. R., Gray, S. E., Lane, T. J., Beck, D. & Collie, A. Workers’ Compensation Claims in Government Employees. https://www.monash.edu/__data/assets/pdf_file/0004/1617277/COMPARE_Government-employees_with-DOI.pdf (2018) doi:10.26180/5c3547ca72b0b.

54. Berecki-Gisolf, J., Clay, F. J., Collie, A. & McClure, R. J. The impact of aging on work disability and return to work. J. Occup. Environ. Med. 54, 318–327 (2012).

55. Krause, N., Dasinger, L. K., Deegan, L. J., Brand, R. J. & Rudolph, L. Alternative approaches for measuring duration of work disability after low back injury based on administrative workers’ compensation data. Am. J. Ind. Med. 35, 604–618 (1999).

56. Collie, A., Di Donato, M. & Iles, R. A. Work disability in Australia: an overview of prevalence, expenditure, support systems and services. J. Occup. Rehabil. 29, 526–539 (2019).

